# Cross-sectional seroprevalence study of Lassa fever in three southern Nigerian states

**DOI:** 10.64898/2025.12.17.25342407

**Authors:** David Simons, Christina Harden, Natalie Imirzian, Katharine Thompson, Nzube Michael Ifebueme, Sunday Eziechina, Helen Ezigbo, Diana Marcus, Fisayomi Aderibigbe, James Koninga, Martin Meremikwu, Lina Moses, David W. Redding, Sagan Friant

## Abstract

Lassa virus (LASV) transmission varies markedly across West Africa, yet its local drivers are poorly understood. We conducted a cross-sectional study in nine villages across three Nigerian states, testing 1,874 individuals for LASV IgG antibodies and analyzing questionnaire data for correlates of exposure. Overall seroprevalence was 3.3% but was highly heterogeneous, ranging from 1.6% to 5.2% between states. We found no consistent demographic, environmental, or behavioural risk factors, and age-seroprevalence patterns differed markedly between villages. Furthermore, we observed no significant village-wide spatial clustering of seropositive households. Our findings indicate that LASV exposure is driven by complex, hyper-local factors rather than by generalizable risk profiles or geography. This challenges the efficacy of spatially-targeted interventions, supporting instead household-level mitigation strategies and integrated One Health approaches to control this endemic disease.

**Article Summary Line:** Lassa virus seroprevalence in rural Nigeria exhibits marked village-level heterogeneity, indicating that transmission is driven by hyper-local, household-scale dynamics rather than broad regional risk factors.

## Introduction

Lassa virus (*Mammarenavirus lassaense*; Arenaviridae, LASV) is a zoonotic arenavirus endemic to West Africa, with regular outbreaks occurring in Nigeria, Guinea, Liberia, and Sierra Leone [1]. It causes Lassa fever, a viral haemorrhagic illness responsible for an estimated 900,000 annual infections and over 200 deaths annually in Nigeria alone [2,3]. Clinical presentation ranges from asymptomatic infection to severe haemorrhagic fever with case fatality exceeding 20% in hospitalized patients, for whom treatment options remain limited [4,5].

The primary reservoir for LASV is *Mastomys natalensis*, a highly fecund, synanthropic rodent widely distributed across sub-Saharan Africa [6]. It thrives in agricultural and peri-domestic environments, creating substantial opportunities for human contact [7]. Infected rodents, particularly those infected vertically in utero, can become persistent shedders of the virus in excreta, while its fluctuating population dynamics influence seasonal transmission intensity [8,9].

Humans are expected to become infected through direct or indirect contact with infected rodents, their excreta, contaminated food or aerosolized particles [10]. Occupational and domestic exposures, particularly in farming, food storage, and rodent hunting contexts, are commonly implicated. Critically, seroprevalence surveys show that exposure is widespread, with most infections (~80%) being asymptomatic or subclinical, meaning case counts severely underestimate the true infection burden [11,12].

Environmental and ecological conditions strongly influence LASV spillover [13]. Land use changes like deforestation and agricultural intensification can increase human-rodent contact, while shifts in rodent community composition may also moderate risk [14–16]. Despite these broad-scale drivers, observed variation in LASV prevalence between ecologically similar areas highlights the critical importance of fine-scale factors [17]. These include social conditions, such as household structure, food storage methods, and specific human-animal interactions.

Current Lassa fever surveillance relies primarily on passive case detection at sentinel hospitals, which overlooks mild infections and asymptomatic seroconversion in the community [18]. Consequently, major gaps remain in understanding why transmission risk varies so markedly at local scales, where conditions facilitating spillover are poorly defined [19]. To address this, we conducted a cross-sectional study in nine communities across three Nigerian states. We combined serological testing with structured questionnaires to investigate fine-scale drivers of exposure. Our objectives were to: 1) estimate LASV seroprevalence at state and village levels; 2) characterize individual and household correlates of seropositivity; and 3) explore spatial heterogeneity in local transmission risk.

## Materials and Methods

The primary objective of this study was to estimate the seroprevalence of LASV IgG in nine communities across three Nigerian states. Secondary objectives were to explore demographic, environmental, behavioural, and occupational correlates of seropositivity and to assess fine-scale spatial heterogeneity of exposure.

### Study design and participants

This cross-sectional baseline survey was conducted as part of the SCAPES longitudinal study [19]. Nine villages across three states (Benue [BN], Ebonyi [EB], and Cross River [CR]) were selected using a tool incorporating population estimates, land cover (as a proxy for *M. natalensis* habitat suitability), and logistical accessibility [20]. Final selection was confirmed through site visits and consultation with local public health surveillance teams.

Households were enumerated to estimate population size and plan recruitment. Villages ranged from 110 to 321 households (population 465–2,313; Appendix Figure 1). We aimed to recruit ~20% of households per village (target ~70), enrolling four individuals per household: an adult male, adult female, adolescent (12–18), and a randomly selected child (<12). Children under 12 provided dried blood spot (DBS) samples but did not complete questionnaires.

Based on published LASV seroprevalence data [17,18,21,22], we assumed high-risk (>40% seroprevalence), medium-risk (10–20%), and low-risk (<5%) village categories. We estimated that 180 individuals would provide 80% power (α = 0.05, β = 0.2) to detect significant differences in LASV seroprevalence between village categories. Household-level clustering of serostatus was expected; hence, the design was powered to detect differences at both village and household levels.

Systematic sampling was used to select households within each village. Routes were designed to approach every *n*th household (where *n* is the total number of households divided by the target sample size). Three households (all in BN) declined participation; in these instances, the next household was enrolled. The participant flow diagram is available in Appendix Figure 2.

### Ethics approval

The study protocol was approved by the Institutional Review Board of the Pennsylvania State University, USA (STUDY00019989), and by the Nigerian National Health Research Ethics Committee (AEC/03/168/24). Additional approval was obtained from the health research ethics committees of the public health agencies in each participating state. Enrollment involved engagement with community leaders. Informed consent was obtained from all adults; assent and parental consent were obtained for minors.

### Data collection

A questionnaire covering demographic, environmental, behavioural, and occupational domains was administered [27]. DBS samples were collected by fingerstick on Whatman 903 cards, air-dried, and transported with desiccant. Samples were eluted and tested for LASV IgG using the Panadea LASV ELISA kit following the manufacturer’s protocol modified for DBS [23,24].

### Statistical methods

Analyses were performed in R (v4.2.3) [25]. Questionnaire responses were linked to individual and household records using unique identifiers. Household GPS coordinates were used for spatial analyses. Data cleaning involved range validation, logical consistency checks, and outlier flagging. Missingness was negligible (<1%); analyses were restricted to complete cases.

#### Descriptive group comparisons

To compare characteristics between villages or states, we fitted separate Bayesian models using the brms package [26] (Gaussian linear models for continuous variables; multinomial logistic regression for categorical). We defined a notable difference between groups as evidence that the variability of the outcome between villages/states was non-zero. This was assessed by examining the posterior distribution of the random effect standard deviation (SD) parameter for the grouping variable. A notable difference was concluded if the 95% Credible Interval (CrI) for the SD parameter excluded zero.

#### Estimating LASV seroprevalence

We estimated seroprevalence and 95% CrIs using Bayesian generalised linear mixed models (GLMM) with a binary outcome (seropositive/seronegative) and a single fixed effect for state or village. Posterior predictions generated marginal estimates averaged over sampled individuals. To explore age-related variation, we fitted a Bayesian generalised additive model (GAM) with village-specific smooth terms for age. Posterior expected probabilities were calculated across a grid of age values to obtain modelled estimates. The full posterior summaries (central estimates and CrIs) for these model comparisons are presented in Appendix Table 1. Differences classed as important are indicated in the main text and tables.

#### Characterising behavioural and exposure correlates of seropositivity

We investigated associations between LASV seropositivity and a range of *a priori* risk factors using a Bayesian GLMM framework, specifying a Bernoulli (logit) likelihood for serostatus as the binary outcome. Analyses were conducted at both individual and household levels, with explanatory variables grouped into four conceptual domains: demographic, environmental, behavioural, and occupational.

To account for potential non-independence of observations within villages, reflecting shared environmental, infrastructural, or cultural exposures, a random intercept for village was included in all models. Univariable models were fitted separately for each covariate to examine associations in isolation, adjusting only for village-level clustering. We evaluated the magnitude and uncertainty of potential risk factors by examining the posterior odds ratios (OR) and their 95% CrIs. An interval that included 1.0 was interpreted as showing no strong evidence for an association.

#### Exploring spatial heterogeneity in LASV exposure

To assess spatial patterns in LASV exposure at the household level, we geocoded each sampled household and identified those with at least one seropositive individual. We first applied Global Moran’s I to test for overall spatial autocorrelation of household seropositivity within each village. To identify potential micro-scale clustering, we then used local Getis-Ord Gi* analysis using the spdep package, which compares the value at each household (the proportion of seropositive members) to the values of neighbouring households within a defined spatial lag (300m), generating a z-score indicating statistically significant hot spots (high-value clusters) and cold spots (low-value clusters) [28].

As a *post hoc* check on the relationship between hotspot classification and seropositivity, we fit a simplified logistic regression model. This model used household serostatus (presence/absence of seropositive individuals) as the outcome and compared two groups: households located in any Gi□-defined cluster (Hotspot or Coldspot) versus those classified as Not Significant. This allowed us to robustly quantify the association between being in a statistically-defined cluster and the household’s observed seropositivity.

Given the low seroprevalence and relatively small village area (median = 6.7km^2^), these analyses were intended as descriptive and exploratory, with a focus on identifying potential micro-scale heterogeneity rather than formally testing for clustering.

## Results

### Study population

Between 16 December 2023 and 22 July 2024, we enrolled 1,926 individuals (1,874 obtained DBS samples, 1,469 completed questionnaires) from 577 households across nine villages in three Nigerian states (BN, CR, EB). This sample represented 27% of households and 11% of the expected total village population (Table 1, Table 2).

**Table 1.**
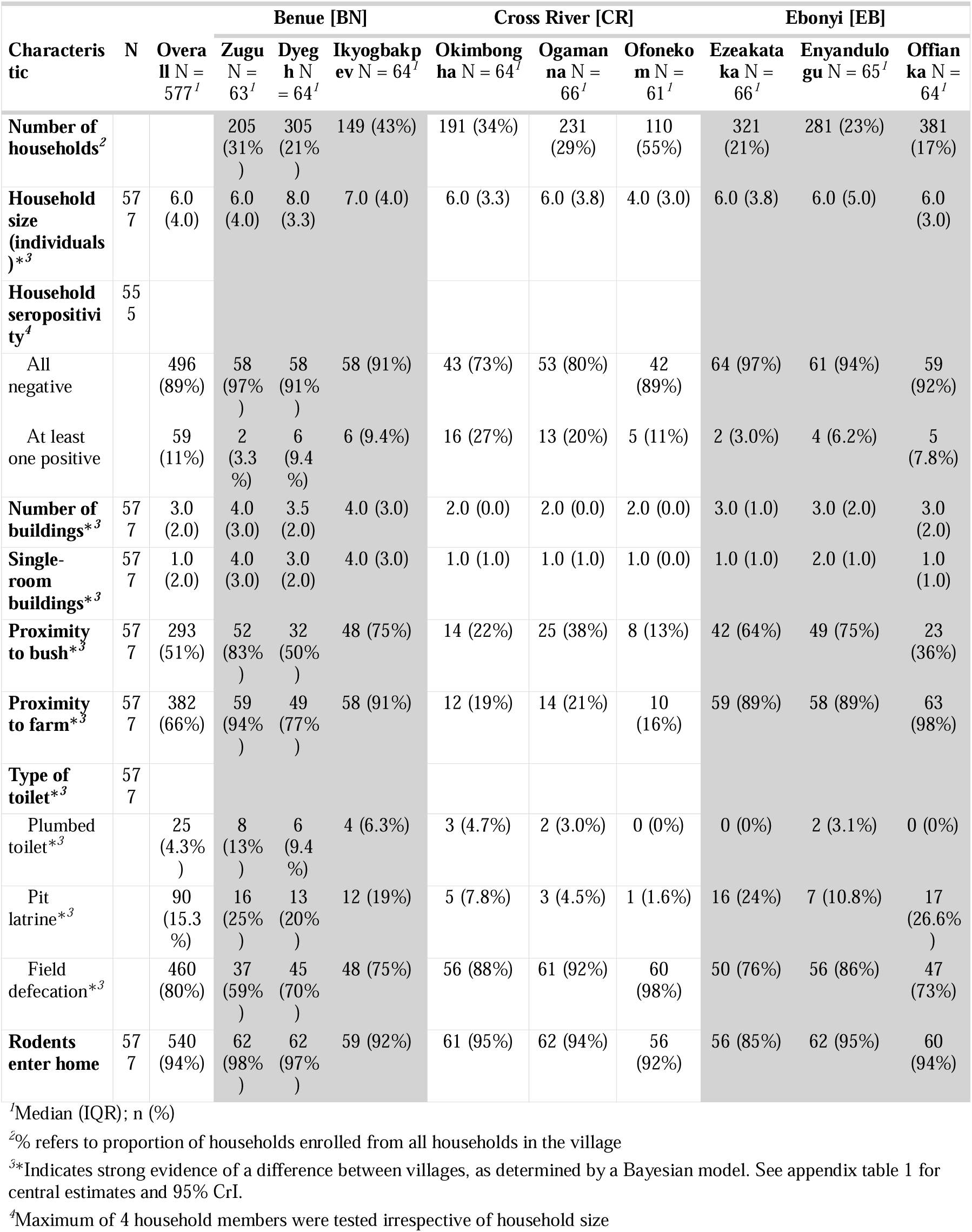
Household-level demographic and environmental characteristics.

**Table 2.**
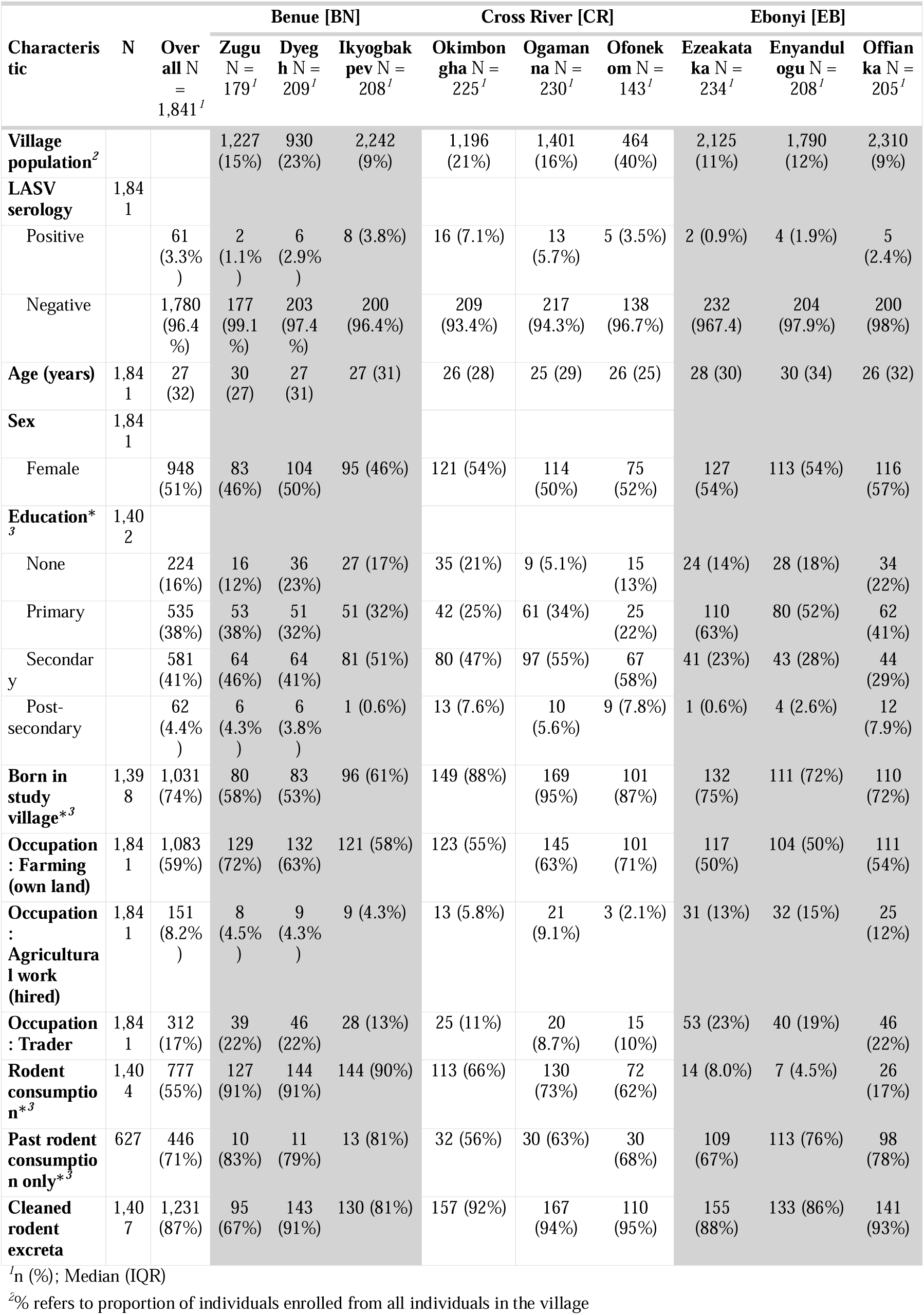
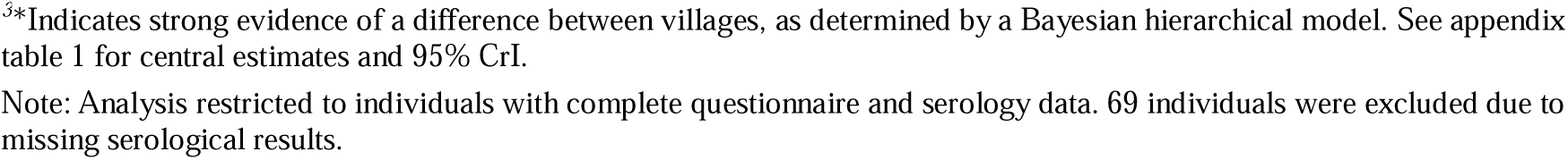
Individual-level demographic, occupational, and behavioral characteristics.

| Characteristic | N | Overall N = 1,841<sup>1</sup> | Benue [BN] |  |  | Cross River [CR] |  |  | Ebonyi [EB] |  |  |
| --- | --- | --- | --- | --- | --- | --- | --- | --- | --- | --- | --- |
|  |  |  | Zugu N = 179<sup>1</sup> | Dyeg h N = 209<sup>1</sup> | Ikyogbak pev N = 208<sup>1</sup> | Okimbon gha N = 225<sup>1</sup> | Ogama na N = 230<sup>1</sup> | Ofonek om N = 143<sup>1</sup> | Ezeakata ka N = 234<sup>1</sup> | Enyandul ugu N = 208<sup>1</sup> | Offian ka N = 205<sup>1</sup> |
| **Village population<sup>2</sup>** |  |  | 1,227 (15%) | 930 (23%) | 2,242 (9%) | 1,196 (21%) | 1,401 (16%) | 464 (40%) | 2,125 (11%) | 1,790 (12%) | 2,310 (9%) |
| **LASV serology** | 1,841 |  |  |  |  |  |  |  |  |  |  |
| Positive |  | 61 (3.3%) | 2 (1.1%) | 6 (2.9%) | 8 (3.8%) | 16 (7.1%) | 13 (5.7%) | 5 (3.5%) | 2 (0.9%) | 4 (1.9%) | 5 (2.4%) |
| Negative |  | 1,780 (96.4%) | 177 (99.1%) | 203 (97.4%) | 200 (96.4%) | 209 (93.4%) | 217 (94.3%) | 138 (96.7%) | 232 (967.4) | 204 (97.9%) | 200 (98%) |
| **Age (years)** | 1,841 | 27 (32) | 30 (27) | 27 (31) | 27 (31) | 26 (28) | 25 (29) | 26 (25) | 28 (30) | 30 (34) | 26 (32) |
| **Sex** | 1,841 |  |  |  |  |  |  |  |  |  |  |
| Female |  | 948 (51%) | 83 (46%) | 104 (50%) | 95 (46%) | 121 (54%) | 114 (50%) | 75 (52%) | 127 (54%) | 113 (54%) | 116 (57%) |
| **Education\*<sup>3</sup>** | 1,402 |  |  |  |  |  |  |  |  |  |  |
| None |  | 224 (16%) | 16 (12%) | 36 (23%) | 27 (17%) | 35 (21%) | 9 (5.1%) | 15 (13%) | 24 (14%) | 28 (18%) | 34 (22%) |
| Primary |  | 535 (38%) | 53 (38%) | 51 (32%) | 51 (32%) | 42 (25%) | 61 (34%) | 25 (22%) | 110 (63%) | 80 (52%) | 62 (41%) |
| Secondary |  | 581 (41%) | 64 (46%) | 64 (41%) | 81 (51%) | 80 (47%) | 97 (55%) | 67 (58%) | 41 (23%) | 43 (28%) | 44 (29%) |
| Post-secondary |  | 62 (4.4%) | 6 (4.3%) | 6 (3.8%) | 1 (0.6%) | 13 (7.6%) | 10 (5.6%) | 9 (7.8%) | 1 (0.6%) | 4 (2.6%) | 12 (7.9%) |
| **Born in study village\*<sup>3</sup>** | 1,398 | 1,031 (74%) | 80 (58%) | 83 (53%) | 96 (61%) | 149 (88%) | 169 (95%) | 101 (87%) | 132 (75%) | 111 (72%) | 110 (72%) |
| **Occupation : Farming (own land)** | 1,841 | 1,083 (59%) | 129 (72%) | 132 (63%) | 121 (58%) | 123 (55%) | 145 (63%) | 101 (71%) | 117 (50%) | 104 (50%) | 111 (54%) |
| **Occupation : Agricultural work (hired)** | 1,841 | 151 (8.2%) | 8 (4.5%) | 9 (4.3%) | 9 (4.3%) | 13 (5.8%) | 21 (9.1%) | 3 (2.1%) | 31 (13%) | 32 (15%) | 25 (12%) |
| **Occupation : Trader** | 1,841 | 312 (17%) | 39 (22%) | 46 (22%) | 28 (13%) | 25 (11%) | 20 (8.7%) | 15 (10%) | 53 (23%) | 40 (19%) | 46 (22%) |
| **Rodent consumption\*<sup>3</sup>** | 1,404 | 777 (55%) | 127 (91%) | 144 (91%) | 144 (90%) | 113 (66%) | 130 (73%) | 72 (62%) | 14 (8.0%) | 7 (4.5%) | 26 (17%) |
| **Past rodent consumption only\*<sup>3</sup>** | 627 | 446 (71%) | 10 (83%) | 11 (79%) | 13 (81%) | 32 (56%) | 30 (63%) | 30 (68%) | 109 (67%) | 113 (76%) | 98 (78%) |
| **Cleaned rodent excreta** | 1,407 | 1,231 (87%) | 95 (67%) | 143 (91%) | 130 (81%) | 157 (92%) | 167 (94%) | 110 (95%) | 155 (88%) | 133 (86%) | 141 (93%) |

<sup>1</sup>n (%); Median (IQR)<sup>2</sup>% refers to proportion of individuals enrolled from all individuals in the village
<sup>3</sup>\*Indicates strong evidence of a difference between villages, as determined by a Bayesian hierarchical model. See appendix table 1 for central estimates and 95% CrI.
Note: Analysis restricted to individuals with complete questionnaire and serology data. 69 individuals were excluded due to missing serological results.

### Households

Household composition and infrastructure showed notable variation across the study villages, particularly in household structure (Table 1). There was strong evidence of differences in household size, ranging from median sizes of 8 in Dyegh [BN] to 4 in Ofonekom [CR]. Similarly, the number of buildings per household and number of single-room buildings differed substantially between villages, with households in BN generally being larger and comprising more buildings.

Environmental characteristics were more consistent across sites. Proximity to bush (51%) and farms (66%), predominant sanitation method (field defecation: 80%), and rodent entry into homes (94%) did not show strong evidence of differing between villages.

Households employed various methods for rodent removal. In contrast to the uniformity of rodent entry, our analysis found strong evidence that the specific methods used, such as poison (73%), sticks (44%), and traps, varied substantially between villages. However, the ultimate outcome for captured rodents was largely consistent; 88% of households reported disposing of them. While disposal was uniform, the practice of consuming captured rodents did show notable variation between villages, consistent with the individual-level data (Table 2).

### Individuals

Demographics and behaviors varied notably across sites (Table 2). Migration patterns differed (Strong Evidence); the percentage of residents born in their current village was substantially lower in BN compared to CR. However, most participants were permanent residents (median 12 months/year). A balanced distribution by sex was achieved by the sampling design for all sites. Median age (Overall: 28, IQR: 31) was consistent across villages.

Educational attainment varied substantialy; participants in BN and CR had higher rates of secondary education completion compared to EB. Subsistence farming was the most common occupation (59%), followed by trading (17%) and hired agricultural work (6%). Primary occupations did not show strong evidence of difference between villages.

Rodent consumption practices differed substantially. Current rodent consumption was common in most BN and CR villages (e.g., 91% in Dyegh [BN] and 73% in Ogamanna [CR]) but was rarer in the EB villages (e.g., 4.5% in Enyandulogu). Even among those not currently consuming rodents, the proportion who reported past consumption also varied, being generally higher in EB villages (e.g., 78% in Offianka) than elsewhere. Indirect rodent contact, such as cleaning rodent excreta, was nearly universal (99%) and did not differ between villages.

### LASV seroprevalence estimates

Of the 1,874 individuals tested, 61 were seropositive for LASV IgG, yielding a crude overall seroprevalence of 3.3%. These individuals belonged to 59 households, with only two households (both in Ikyogbakpev [BN]) containing more than one seropositive member. No seropositive individuals reported a prior diagnosis of Lassa fever.

Model-based estimates revealed marked heterogeneity in exposure risk across the study area (Table 3). The overall seroprevalence was 3.17% (95% CrI: 2.47-4.04%). This varied at the state level (CR = 5.13% [3.66–6.97%]; BN = 2.63% [1.56–4.06%]; EB = 1.65% [0.86–2.85%]) and showed even greater variation at the village level (ranging from Ezeakataka [EB] = 0.79% [0.13-2.45%] to Okimbongha [CR] = 6.48% [3.9-9.97%]) (Table 3.).

**Table 3.** Model-based LASV IgG seroprevalence estimates at the state and village level.

| State/Village | N sampled | N positive | Prevalence (%) | 95% CrI |
| --- | --- | --- | --- | --- |
| **Benue** | 616 | 16 | 2.63 | 1.56 - 4.06 |
| Dyegh | 216 | 6 | 2.74 | 1.08 - 5.47 |
| Ikyogbakpev | 214 | 8 | 3.72 | 1.7 - 6.82 |
| Zugu | 186 | 2 | 1.03 | 0.17 - 3.28 |
| **Cross River** | 597 | 34 | 5.13 | 3.66 - 6.97 |
| Ofonekom | 140 | 5 | 2.67 | 0.96 - 5.73 |
| Ogamanna | 234 | 13 | 5.53 | 3.06 - 8.94 |
| Okimbongha | 223 | 16 | 6.48 | 3.9 - 9.97 |
| **Ebonyi** | 661 | 11 | 1.65 | 0.86 - 2.85 |
| Enyandulogu | 215 | 4 | 1.81 | 0.55 - 4.23 |
| Ezeakataka | 239 | 2 | 0.79 | 0.13 - 2.45 |
| Offianka | 207 | 5 | 2.31 | 0.83 - 4.88 |

The relationship between age and seropositivity was not uniform, revealing distinct, village-specific transmission dynamics (Figure 1). Two villages (Ogamanna [CR] and Ofonekom [CR]) exhibited the classic pattern of increasing seroprevalence with age, consistent with long-term, cumulative exposure. This trend was statistically robust in Ogamanna [CR] (Posterior probability of increase > 99%), while Ofonekom [CR] showed a similar but more uncertain trajectory (Probability ~76%). Notably, these two villages are geographically adjacent (< 5km apart), suggesting a shared local ecology. Conversely, Okimbongha [CR] and Ezeakataka [EB] displayed profiles peaking in younger groups before declining, suggestive of recent, focal exposure, though statistical evidence for the decline was weaker (Probability 60–66%). In remaining villages, seroprevalence remained consistently low.

**Fig 1.**
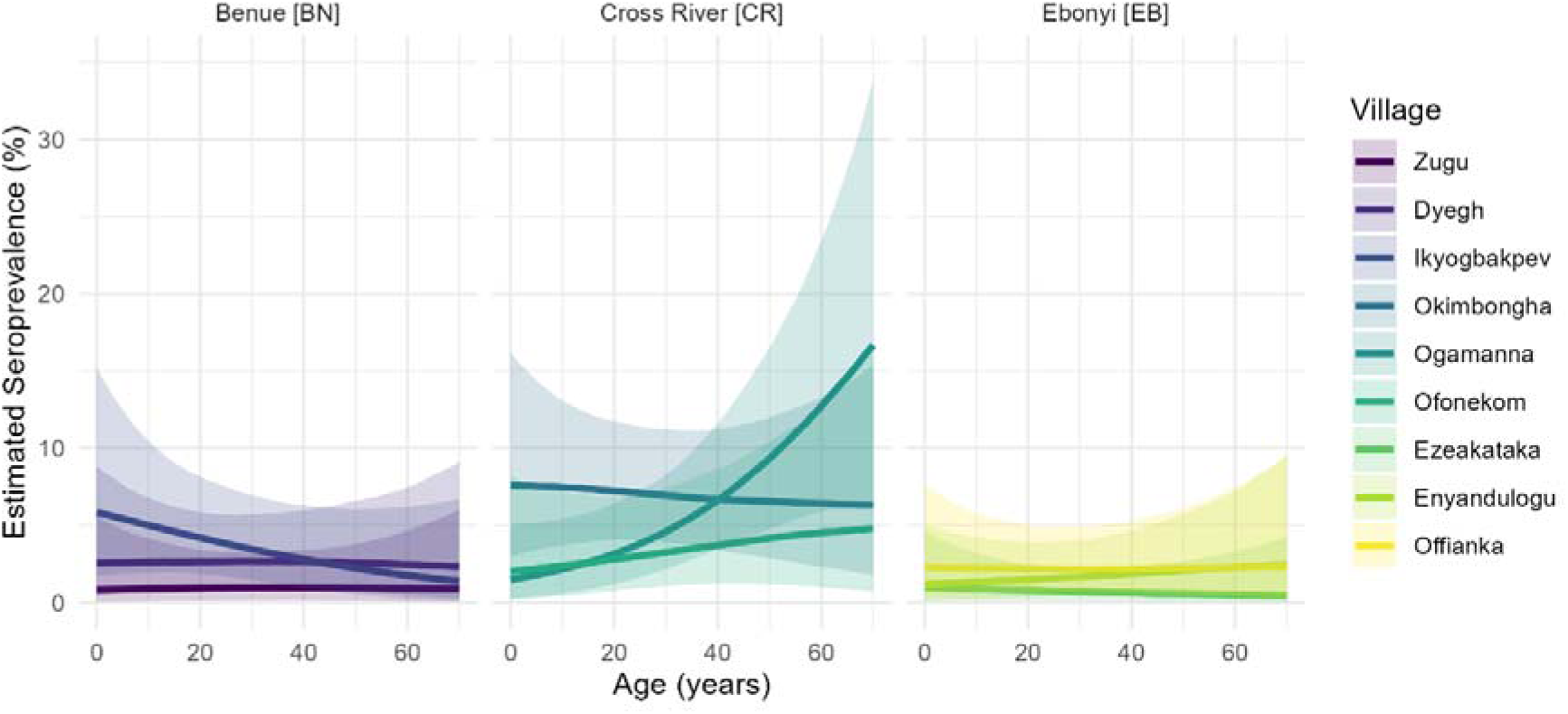
Village-specific age-seroprevalence curves for Lassa virus. Lines represent median posterior estimates from a generalized additive model, and ribbons show the 95% credible interval.

### Risk factors for seropositivity

Univariable models adjusting for village-level clustering found no strong evidence of association between seropositivity and most pre-specified risk factors (Figure 2). Odds ratios were generally close to 1.0 with wide intervals. Given these results and the study’s power limitations from overall low seropositivity, multivariable modelling was deemed unnecessary, we therefore only report univariable results below.

**Fig 2.**
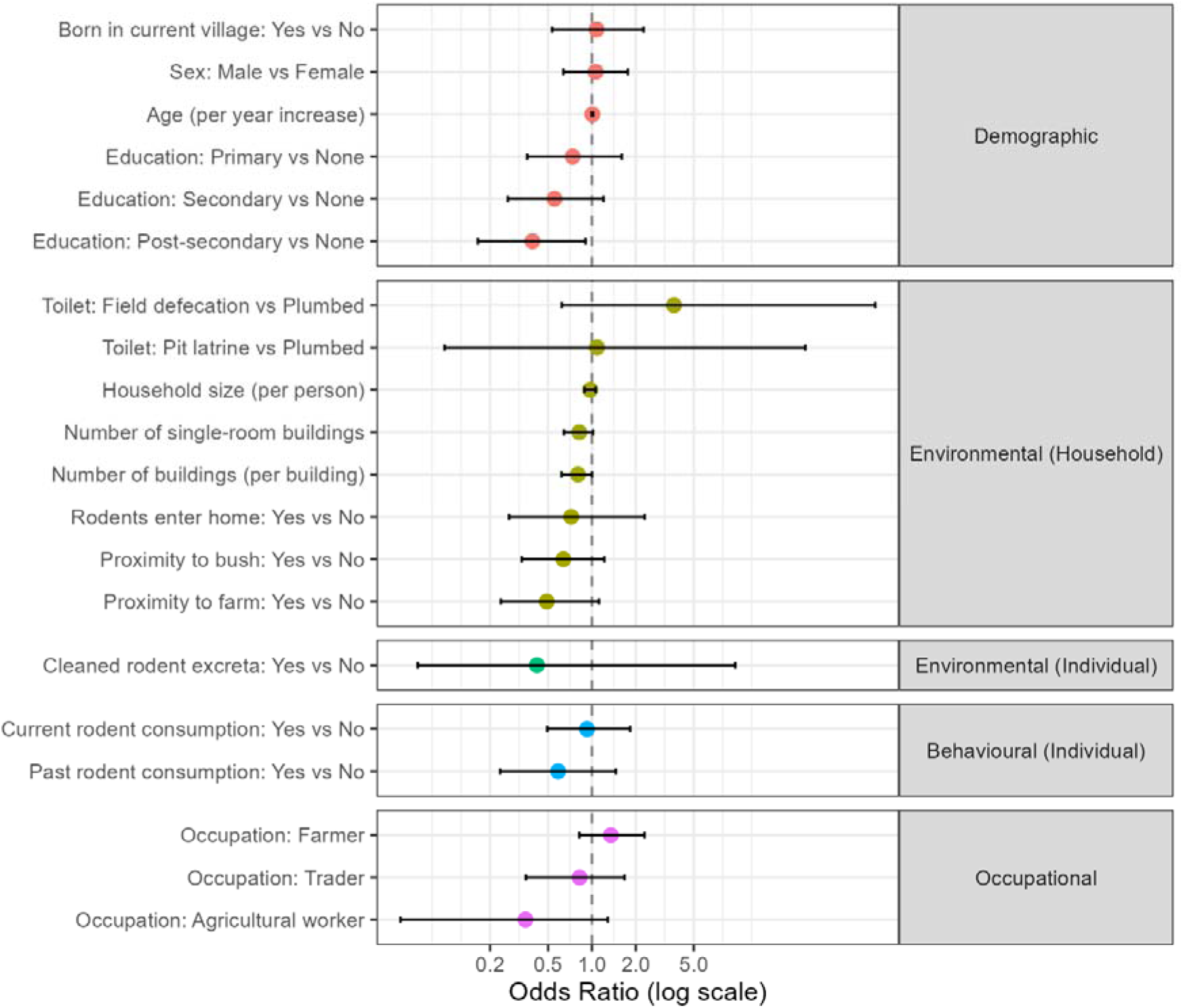
Forest plot of univariable odds ratios for Lassa virus seropositivity. Points are posterior medians, and horizontal bars are 95% credible intervals, adjusted for clustering by village.

#### Demographic correlates of LASV seropositivity

No meaningful association was found for age (OR: 1.01 [0.99–1.02]) or sex (OR: 1.06 [0.64–1.76]). Post-secondary education showed a protective trend (OR: 0.39 [0.17–0.90]), though likely confounded by age and socioeconomic status.

#### Environmental and Behavioral correlates of LASV seropositivty

No clear associations were found for household-level variables such as household size (OR: 0.97, [0.89–1.06]), number of buildings (OR: 0.80, [0.62–1.00]), rodent entry into the home (OR: 0.72, [0.27–2.30]), or cleaning of rodent excreta (OR: 0.42 [0.06–9.64]). Odds ratios for rodent consumption (OR: 0.93 [0.49–1.83]) and past rodent consumption only (OR: 0.59 [0.23–1.46]) were also uncertain.

### Spatial heterogeneity in LASV exposure

We assessed the spatial clustering of LASV seropositivity at the household level across nine villages. Global spatial autocorrelation, assessed using Moran’s I, found no evidence of significant village-wide clustering in any site (all *p* > 0.05). Localised clustering was then evaluated using the Getis-Ord Gi* statistic, which identified a total of 27 hotspot and 6 coldspot households across all villages (Figure 3). However, these statistical clusters did not align well with observed seropositivity; of the 59 seropositive households in the study, only 3.6% were located within a designated hotspot.

**Fig 3.**
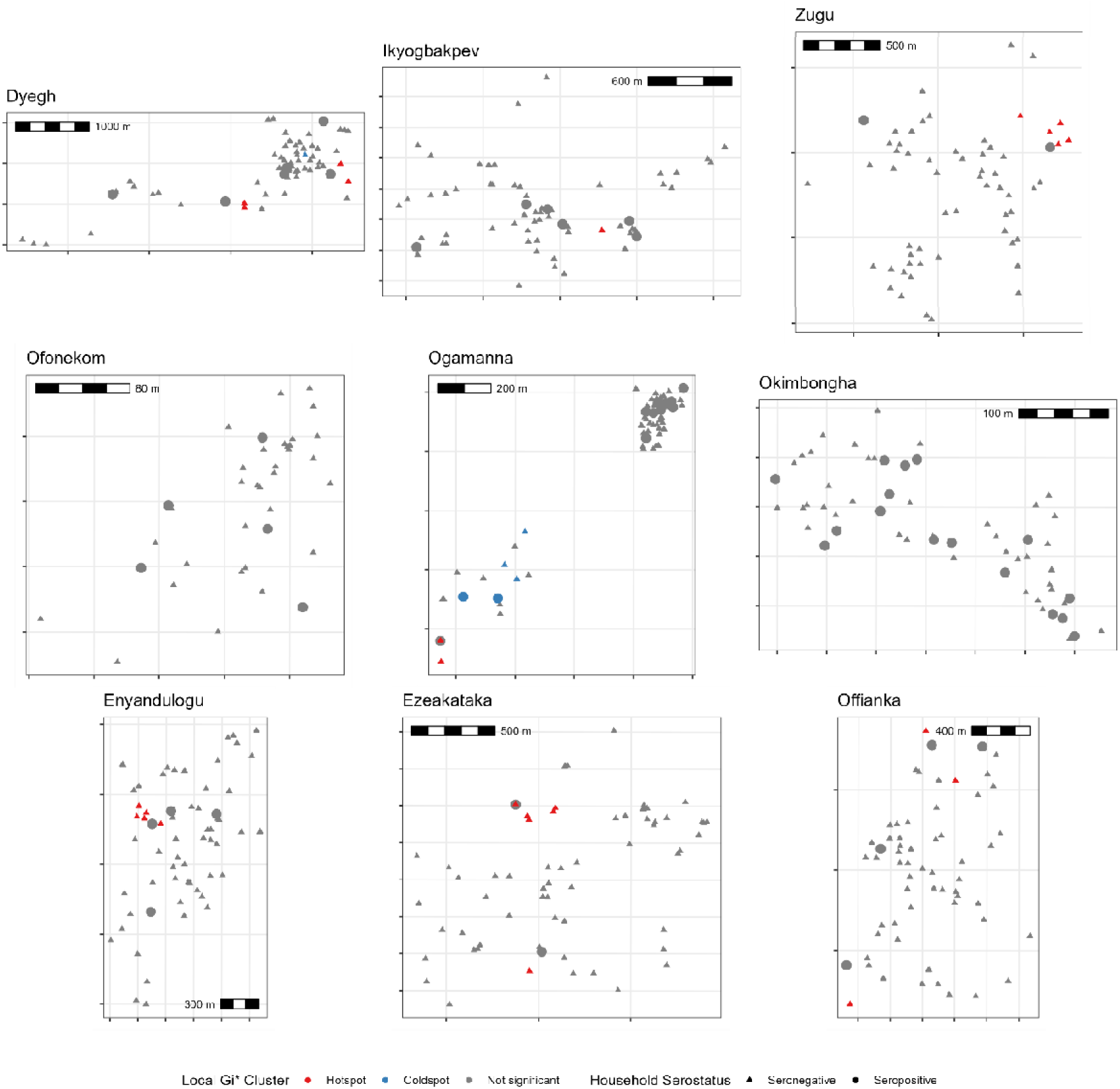
Spatial distribution of household LASV seropositivity and Local Gi* clustering. Each panel represents a single village, showing jittered household locations. Shapes indicate whether at least one LASV-seropositive individual was recorded in the household. The color classification reflects the local clustering of seropositivity, where the Gi* test includes the household’s own value in the local average; a high-value central household may thus be classified as ‘Not significant’ if surrounding seronegative households dilute its local average below the statistical threshold. Scale bars are included to indicate spatial scale; coordinates are not shown to preserve privacy.

Logistic regression confirmed no significant difference in the odds of seropositivity based on cluster status (*p* = 0.39 *OR_clustered vs. non-clustered_* = 0.53 [0.12-2.3]). These results strongly indicate that, in our study sites, seropositive households were not concentrated in discernible spatial clusters.

## Discussion

This study found that LASV exposure in rural Nigeria was substantially lower than anticipated based on national risk models [13]. While the overall model-based IgG seroprevalence was 3.2% [2.5–4.0%], this aggregate figure masks marked heterogeneity at the local scale. Prevalence varied significantly between states (1.7% to 5.1%) and, more critically, between villages (0.8% to 6.5%). These findings challenge broad-scale risk mapping, pointing to a transmission landscape that is generally low but punctuated by complex, hyper-local variation supported by three observations: divergent age patterns, absence of risk factor correlation, and lack of spatial clustering.

Our seroprevalence estimates are comparable to those from some contemporary studies in West Africa, though such comparisons are conditioned by differences in study populations and laboratory methods [17,29]. The high specificity of the commercial ELISA kit (LASV vs. non-LASV arenaviruses) used in this study may yield more conservative estimates than alternative serological assays [24]. More revealing was the relationship with age. Two villages exhibited classic increasing seroprevalence with age, indicative of cumulative, endemic transmission. Conversely, two others showed peaks in younger groups, suggesting dynamic local transmission [30].

Peaks in younger age groups could arise from recent increases in transmission or immunological factors like seroreversion. With antibody waning estimated at ~3% annually [31], the decline in older cohorts likely reflects the loss of detectable markers from historical infections. This implies that cross-sectional seroprevalence may serve only as a moving window of recent exposure (e.g., the last 15–20 years) rather than a cumulative record of lifetime risk. Consequently, the distinct peak in younger age groups observed in villages like Okimbongha [CR] likely signals intense, recent transmission events concentrated in younger cohorts, while the low prevalence in older adults reflects both lower recent exposure and the decay of historical antibodies.

Consistent with this theme of local heterogeneity, our univariable analyses revealed no strong or consistent demographic, environmental, or occupational correlates of seropositivity. The lack of clear associations with commonly cited risks, such as farming, contrasts with some previous findings [32]. This may be a function of the study’s limited statistical power, a consequence of the low overall seroprevalence, to detect modest individual-level effects. Furthermore, the near-universal reporting of certain exposures, such as the presence of rodents in homes (94%), creates little variability. This complicates the quantification of their true impact, particularly as risk is ultimately driven by the prevalence of LASV within local rodent populations, which itself can be highly variable, again highlighting the heterogeneous nature of risk [7,14].

The lack of association with rodent consumption, despite its high prevalence in BN and CR (up to 91%), challenges the assumption that hunting or consumption is the primary driver of spillover. It suggests that transmission may occur more frequently through indirect routes, such as contamination of food stores or household surfaces by rodent urine, which affects the entire household regardless of who consumes the meat

Collectively, these findings suggest a departure from a uniform transmission model, indicating instead a process that is intensely local and patchy. It is plausible that LASV exposure risk is governed by a complex interplay of factors at the micro-scale of the household or individual, rather than being broadly structured by village geography or demographics. Such factors may include unmeasured variables like household-specific rodent density and viral shedding status, subtle differences in food storage and sanitation practices, or individual activities in high-risk locations not fully captured by our questionnaire.

### Strengths and Limitations

The strengths of this study include its systematic, multi-criteria site selection; a large sample size across diverse communities; and a robust analytical framework using Bayesian mixed models to account for data hierarchies and uncertainty. The inclusion of formal spatial statistics adds rigor to our conclusion that transmission is not strongly clustered at the household level. Nevertheless, the study has important limitations. It was powered to detect village-level differences based on higher seroprevalence estimates from previous studies; however, the observed low seroprevalence limited our power for individual-level risk factor analysis. The cross-sectional design precludes assessment of causality and the distinction between recent and past infections. This challenge is compounded by the potential for antibody waning and sero-reversion, the rates of which are critical for accurate Force of Infection estimation but remain poorly understood for LASV. Furthermore, the measurement of behaviour and environment at a single cross-sectional point may not accurately reflect cumulative lifetime exposure, as housing and habits vary over time. Finally, reliance on self-reported behaviours is subject to recall bias, and the study lacked objective ecological measures, such as rodent abundance data.

### Public Health Implications and Future Directions

Our findings have significant implications for LASV control. The absence of spatial clustering and a clear risk profile suggests geographically-targeted interventions may be inefficient. Public health efforts should prioritize broadly-deployed, household-level interventions (e.g., rodent-proofing) and maintain high suspicion in surveillance across all villages within endemic regions, not just sentinel sites. Surveillance must also include areas of previously low-predicted risk (e.g., cold spots) given the observed micro-heterogeneity. Future research must employ a One Health approach, leveraging data from our ongoing longitudinal rodent sampling [19]. Ultimately, disentangling these patterns requires longitudinal cohort studies linking exposures with seroconversion.

## Conclusion

LASV exposure in rural Nigeria is highly variable and likely driven by complex, hyper-local factors. Effective control requires shifting from universal risk factors toward adaptable, household-focused interventions and integrated surveillance responding to unique local contexts.

## Data Availability

All data produced are available online at https://github.com/RiskLabPSU/baseline-seroprev-public

## Acknowledgments

We sincerely thank the residents and leadership of the nine study communities in Benue, Cross River, and Ebonyi states for their warm welcome, hospitality, and willingness to participate in this research. We are deeply grateful to the community liaisons and village guides whose local knowledge and dedication were vital for facilitating community entry and navigation. We also acknowledge the State Ministries of Health and the local public health teams, including the Disease Surveillance and Notification Officers (DSNOs), for their essential logistical support and ongoing collaboration throughout the fieldwork. Funding was provided by the joint NSF□NIH□NIFA Ecology and Evolution of Infectious Disease Award #2208034 in partnership with Research and Innovation (UKRI) Biotechnology and Biological Sciences Research Council (BBSRC) Award BB/X005364/1. The authors acknowledge the use of Gemini (Google) for assistance with language editing and refining the clarity of the manuscript text.

## Author Biography

Dr. Simons is a medical doctor and post-doctoral researcher investigating the ecology and epidemiology of Lassa fever. Their broader research focuses on understanding the transmission dynamics of rodent-borne zoonoses to inform disease control strategies.

## Appendix Figures

Appendix Figure 1 Estimated study village populations. Total village populations were estimated using a Bayesian hierarchical Gamma regression model based on household size counts from the survey. The distributions (half-violins) show the posterior probability density of the population estimate. Points represent the central estimate (posterior median), and the lines represent the interquartile range (50% credible interval).

Appendix Figure 2 Participant flow diagram. Study villages were selected from a list of populated places in the three study states. Household enumeration was conducted in each selected village, informing sample size calculations. 577 households were enroled with 1,841 individuals providing dried blood spots for LASV serology, 1,420 individuals were asked to complete the individual level questionnaire.

## Appendix Tables

Appendix Table 1 Bayesian analysis of between-village heterogeneity. This table summarizes the posterior distribution of the random effect standard deviation (SD) parameter for each variable of interest. Estimate represents the median of the posterior distribution for the between-village SD, and 95% CrI represents the 95% Credible Interval. Strong Evidence of a difference is declared if the lower bound of the 95% CrI for the SD parameter is >0.1, indicating that the variation between villages is non-negligible. For multinomial variables (e.g., Education), parameters represent the variation in the log-odds of being in a specific category relative to the reference group.

## References

[1] Garry RF. Lassa fever — the road ahead. Nature Reviews Microbiology 2023;21:87–96. 10.1038/s41579-022-00789-8.

[2] Basinski AJ, Fichet-Calvet E, Sjodin AR, Varrelman TJ, Remien CH, Layman NC, et al. Bridging the gap: Using reservoir ecology and human serosurveys to estimate Lassa virus spillover in West Africa. PLOS Computational Biology 2021;17:e1008811. 10.1371/journal.pcbi.1008811.

[3] Nigeria Centre for Disease Control and Prevention. Lassa Fever Outbreak Reports 2025.

[4] Kenmoe S, Tchatchouang S, Ebogo-Belobo JT, Ka’e AC, Mahamat G, Guiamdjo Simo RE, et al. Systematic review and meta-analysis of the epidemiology of Lassa virus in humans, rodents and other mammals in sub-Saharan Africa. PLoS Neglected Tropical Diseases [Electronic Resource] 2020;14:e0008589. 10.1371/journal.pntd.0008589.

[5] Salam AP, Duvignaud A, Jaspard M, Malvy D, Carroll M, Tarning J, et al. Ribavirin for treating Lassa fever: A systematic review of pre-clinical studies and implications for human dosing. PLoS Neglected Tropical Diseases 2022;16:e0010289. 10.1371/journal.pntd.0010289.

[6] Bellocq JGD, Bryjová A, Martynov AA, Lavrenchenko LA. Dhati Welel virus, the missing mammarenavirus of the widespread Mastomys natalensis. Journal of Vertebrate Biology 2020;69:20018.1. 10.25225/jvb.20018.

[7] Mariën J, Iacono GL, Rieger T, Magassouba N, Günther S, Fichet-Calvet E. Households as hotspots of Lassa fever? Assessing the spatial distribution of Lassa virus-infected rodents in rural villages of Guinea. Emerging Microbes & Infections 2020;9:1055–64. 10.1080/22221751.2020.1766381.

[8] Hoffmann C, Krasemann S, Wurr S, Hartmann K, Adam E, Bockholt S, et al. Lassa virus persistence with high viral titers following experimental infection in its natural reservoir host, Mastomys natalensis. Nature Communications 2024;15:9319. 10.1038/s41467-024-53616-4.

[9] Leirs H, Kirkpatrick L, Sluydts V, Sabuni C, Borremans B, Katakweba A, et al. Twenty-nine years of continuous monthly capture-mark-recapture data of multimammate mice (Mastomys natalensis) in Morogoro, Tanzania. Scientific Data 2023;10:798. 10.1038/s41597-023-02700-3.

[10] Bausch DG, Demby AH, Coulibaly M, Kanu J, Goba A, Bah A, et al. Lassa fever in Guinea: I. Epidemiology of human disease and clinical observations. Vector Borne and Zoonotic Diseases (Larchmont, NY) 2001;1:269–81. 10.1089/15303660160025903.

[11] McCormick JB, Webb PA, Krebs JW, Johnson KM, Smith ES. A prospective study of the epidemiology and ecology of lassa fever. Journal of Infectious Diseases 1987;155:437–44.

[12] Simons D. Lassa fever cases suffer from severe underreporting based on reported fatalities. International Health 2022. 10.1093/inthealth/ihac076.

[13] Redding DW, Moses LM, Cunningham AA, Wood J, Jones KE. Environmental-mechanistic modelling of the impact of global change on human zoonotic disease emergence: A case study of Lassa fever. Methods in Ecology and Evolution 2016;7:646–55. 10.1111/2041-210X.12549.

[14] Bangura U, Buanie J, Lamin J, Davis C, Bongo GN, Dawson M, et al. Lassa Virus Circulation in Small Mammal Populations in Bo District, Sierra Leone. BIOLOGY-BASEL 2021;10. 10.3390/biology10010028.

[15] Eskew EA, Bird BH, Ghersi BM, Bangura J, Basinski AJ, Amara E, et al. Reservoir displacement by an invasive rodent reduces Lassa virus zoonotic spillover risk. Nature Communications 2024;15:3589. 10.1038/s41467-024-47991-1.

[16] Simons D, Gibb R, Bangura U, Sondufu D, Lamin J, Koninga J, et al. Land use gradients drive spatial variation in Lassa fever host communities in the Eastern Province of Sierra Leone. 2025. 10.32942/X2N33P.

[17] Grant DS, Engel EJ, Roberts Yerkes N, Kanneh L, Koninga J, Gbakie MA, et al. Seroprevalence of anti-lassa virus IgG antibodies in three districts of sierra leone: A cross-sectional, population-based study. PLoS Negl Trop Dis 2023;17:e0010938.

[18] Mariën J, Nuismer SL, Magassouba N, Soropogui B, Günther S, Becker-Ziaja B, et al. Serosurveillance Identifies an Endemic Hotspot of Lassa Fever in Faranah, Upper Guinea. The Journal of Infectious Diseases 2025. 10.1093/infdis/jiaf308.

[19] Friant S, Simons D, Bjornstad O, Gibb R, Harden C, Imirzian N, et al. Scaling Lassa Virus Dynamics within Anthropogenic Ecosystems (SCAPES) study: A mixed-methods observational cohort study of humans, rodents, and landscapes in Nigeria 2025:2025.09.12.25335176. 10.1101/2025.09.12.25335176.

[20] Imirzian N, Simons D, Harden C, Johnson E, Hewitson M, Friant S, et al. Sitetool: An application for field site selection and evaluation 2025. 10.32942/X2G65X.

[21] Kernéis S, Koivogui L, Magassouba N, Koulemou K, Lewis R, Aplogan A, et al. Prevalence and risk factors of lassa seropositivity in inhabitants of the forest region of guinea: A cross-sectional study. PLoS Negl Trop Dis 2009;3:e548.

[22] Tobin E, Asogun D, Akpede N, Adomeh D, Odia I, Gunther S. Lassa fever in nigeria: Insights into seroprevalence and risk factors in rural edo state: A pilot study. J Med Trop 2015;17:51.

[23] Longet S, Leggio C, Bore JA, Key S, Tipton T, Hall Y, et al. Influence of landscape patterns on exposure to lassa fever virus, guinea. Emerg Infect Dis 2023;29:304–13.

[24] Soubrier H, Bangura U, Hoffmann C, Olayemi A, Adesina AS, Günther S, et al. Detection of lassa virus-reactive IgG antibodies in wild rodents: Validation of a capture enzyme-linked immunological assay. Viruses 2022;14:993.

[25] R Core Team. R: A language and environment for statistical computing. Vienna, Austria: R Foundation for Statistical Computing; 2023.

[26] Bürkner P-C. brms: An R package for Bayesian multilevel models using Stan. Journal of Statistical Software 2017;80:1–28. 10.18637/jss.v080.i01.

[27] RiskLabPSU. Baseline LASV Seroprevalence Public Repository [Internet]. 2024 [cited 2025 Dec 16]. Available from: https://github.com/RiskLabPSU/baseline-seroprev-public

[28] Bivand R, Wong DWS. Comparing implementations of global and local indicators of spatial association. TEST 2018;27:716–48. 10.1007/s11749-018-0599-x.

[29] Lerch A, Ten Bosch QA, L’Azou Jackson M, Bettis AA, Bernuzzi M, Murphy GAV, et al. Projecting vaccine demand and impact for emerging zoonotic pathogens. BMC Medicine 2022;20:202. 10.1186/s12916-022-02405-1.

[30] Hay JA, Routledge I, Takahashi S. Serodynamics: A primer and synthetic review of methods for epidemiological inference using serological data. Epidemics 2024;49:100806. 10.1016/j.epidem.2024.100806.

[31] Moore SM, Rapheal E, Mendoza Guerrero S, Dean NE, Stoddard ST. Estimation of Lassa fever incidence rates in West Africa: Development of a modeling framework to inform vaccine trial design. PLOS Neglected Tropical Diseases 2025;19:e0012751. 10.1371/journal.pntd.0012751.

[32] Ogundele GO, Jolayemi KO, Bello S. Lassa fever in West Africa: A systematic review and meta-analysis of attack rates, case fatality rates and risk factors. BMC Public Health 2025;25:2948. 10.1186/s12889-025-24377-6.

