## Supplementary figures and images for "Cross-sectional seroprevalence study of Lassa fever in three southern Nigerian states"

### Appendix Figure 1

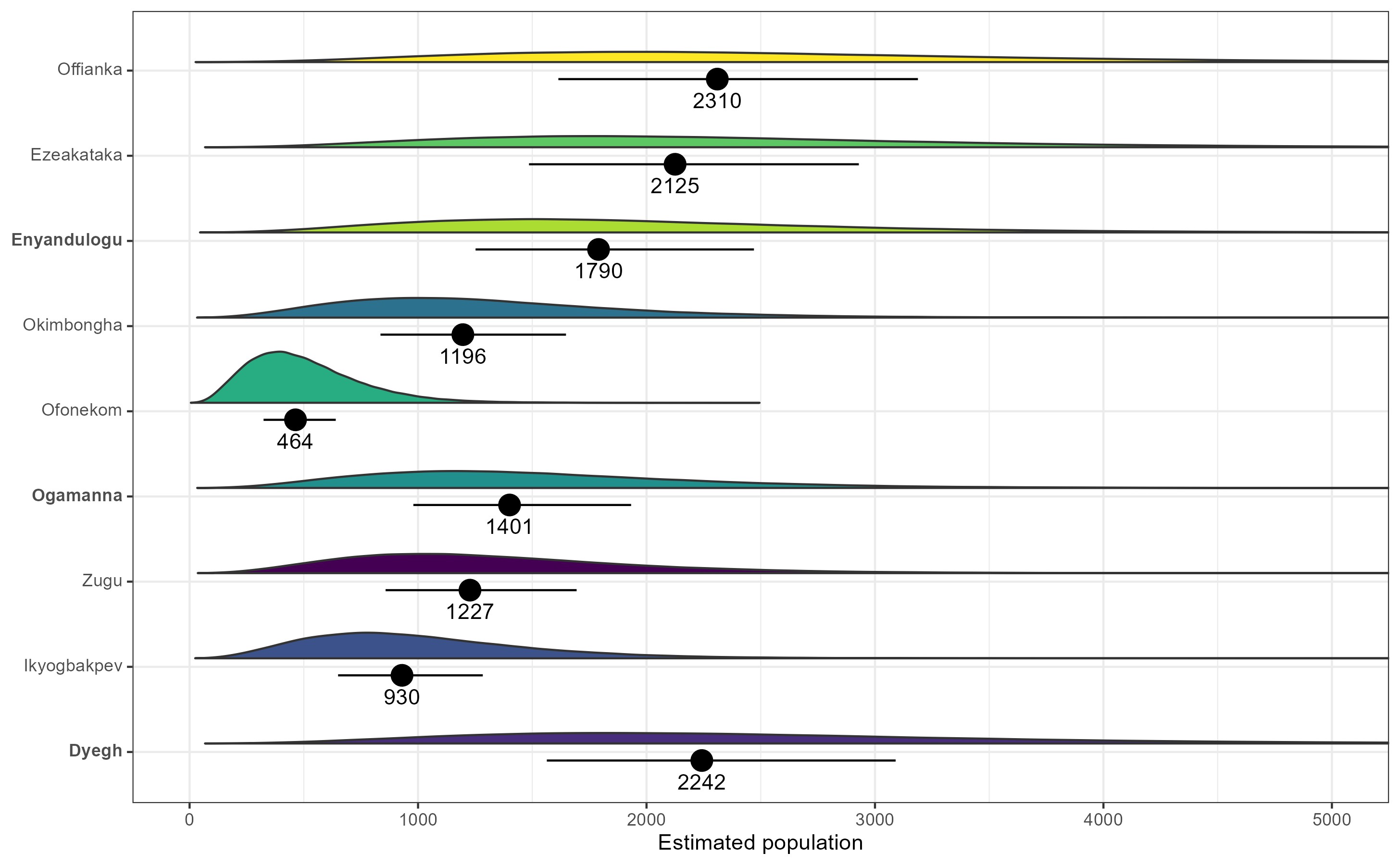

### Appendix Figure 2

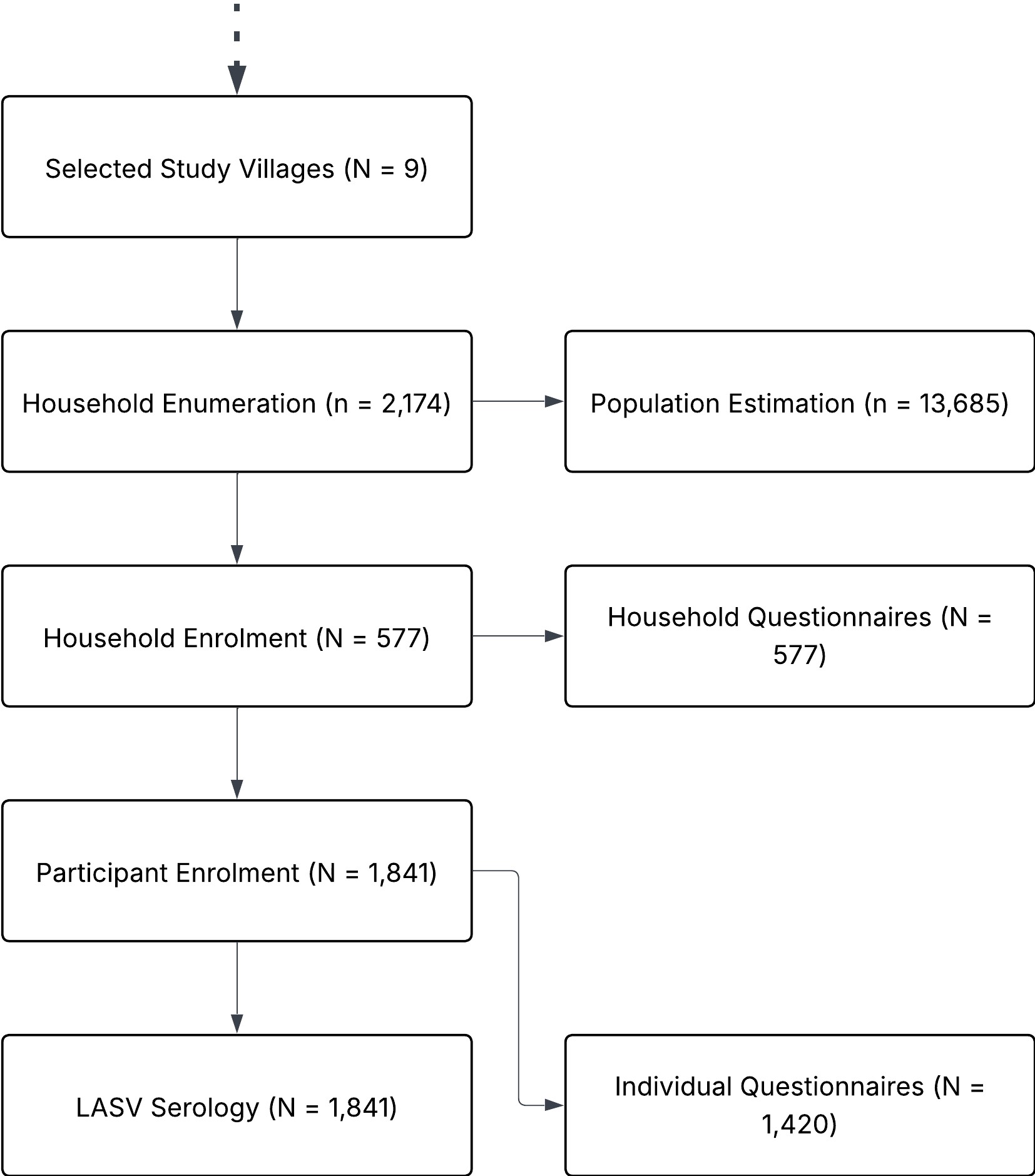
